# Enhanced local synchronization with impaired large-scale recruitment of NREM slow waves in pediatric disorders of arousal

**DOI:** 10.64898/2026.10.13.26365319

**Authors:** Damiana Bergamo, Matteo Pereno, Giovanni Lombardi, Silvia Miano, Salvatore Galati, Giulio Bernardi, Mauro Manconi, Anna Castelnovo

## Abstract

NREM slow waves emerge from the synchronization of neuronal activity across multiple spatial scales. During childhood, local synchronization and large-scale propagation undergo distinct maturational trajectories, paralleling cortical synaptic remodeling and the maturation of long-range brain connectivity. How alterations in this local-to-global organization relate to arousal instability remains poorly understood. Here, we used 256-channel high-density EEG to characterize NREM oscillatory organization in 17 children with disorders of arousal (DOA) and 17 age- and sex-matched healthy controls. Children with DOA showed a marked dissociation between local synchronization and large-scale coordination: slow waves were locally larger, steeper and more synchronized, yet recruited the cortex less globally and exhibited a more spatially restricted distribution of origins. This phenotype preferentially involved Type 1 slow waves, a subtype associated with widespread cortical and subcortico-cortical recruitment and late developmental maturation. Children with DOA also showed reduced centro-posterior slow spindle density, providing convergent evidence for altered coordination of distributed NREM oscillations. These microstructural abnormalities occurred despite largely preserved conventional sleep architecture. Together, our findings reveal an altered local-to-global organization of NREM sleep in pediatric DOA and suggest that vulnerability to incomplete arousals may emerge when strong local synchronization is not matched by effective large-scale network coordination during brain development.

## Introduction

Non-rapid eye movement (NREM) sleep is characterized by slow waves that reflect the synchronization of neuronal activity across multiple spatial scales. Individual slow waves can remain relatively local or recruit large portions of the cortex, depending on the extent and propagation of the underlying neuronal activity (Massimini et al., 2004; Nir et al., 2011). Importantly, the local strength of synchronization and its large-scale spatial recruitment represent partly distinct properties of NREM slow oscillations. Both undergo profound changes during childhood and adolescence, alongside cortical synaptic remodeling and the maturation of long-range structural connectivity (Bergamo et al., 2024; Timofeev et al., 2020). Slow-wave amplitude and slope decline across development in parallel with changes in cortical structure and synaptic organization, whereas the capacity of slow waves to propagate across distant cortical regions increases with the maturation of long-range white-matter connections (Castelnovo et al., 2023; Kurth et al., 2010, 2017). Thus, the developing brain represents a model in which local synchronization and large-scale coordination of NREM activity may become dissociated.

Slow waves are themselves heterogeneous in their spatial organization and putative generating mechanisms (Bergamo et al., 2026; Bernardi et al., 2018; Siclari et al., 2014). Large, steep and highly synchronized Type 1 slow waves are characterized by widespread cortical recruitment and have been associated with arousal-related processes, possibly reflecting the influence of subcortical arousal systems on cortical activity. In contrast, Type 2 waves are typically more local and are thought to predominantly reflect cortico-cortical interactions. Importantly, these slow-wave classes also follow distinct developmental trajectories, with Type 1 waves emerging relatively late during maturation (Castelnovo et al., 2023; Spiess et al., 2018). This combination of widespread recruitment, developmental sensitivity and association with arousal-related mechanisms makes Type 1 slow waves particularly relevant to disorders characterized by incomplete transitions between sleep and wakefulness. Sleep spindles provide a complementary window onto NREM network organization, reflecting coordinated thalamocortical activity and undergoing marked developmental changes in density, frequency and topography (Fernandez & Lüthi, 2020). Together, slow-wave propagation and spindle activity provide complementary markers of distributed NREM coordination.

Disorders of arousal (DOA), including sleepwalking, confusional arousals and sleep terrors, offer a window into the local-to-global organization of NREM sleep. DOA arise predominantly from slow-wave sleep, peak during childhood and frequently disappear with maturation. Their defining neurophysiological feature is incomplete arousal, in which wake-like activation of motor and limbic regions coexists with persistent sleep-like activity in associative cortical areas, indicating a breakdown in the normally coordinated transition between sleep and wakefulness (Bassetti et al., 2000; Castelnovo et al., 2026; Desjardins et al., 2017; Flamand et al., 2018; Januszko et al., 2016; Terzaghi et al., 2009, 2012; Valomon et al., 2026). Yet most physiological studies have focused on adults and on the dynamics immediately preceding or accompanying behavioral episodes. Baseline NREM organization in children with DOA - the developmental period in which the disorder is most prevalent - remains largely unexplored.

Previous studies in adults have reported alterations in slow-wave activity and slow-wave morphology in DOA (Castelnovo et al., 2016), but whether these reflect a generalized change in sleep depth or a more specific alteration in the spatial organization of NREM oscillations remains unclear. This distinction may be particularly important during childhood, when stronger local synchronization need not imply more effective recruitment of distributed cortical networks. We therefore hypothesized that pediatric DOA would be characterized by an altered balance between local synchronization and large-scale NREM coordination, rather than by a uniform increase or decrease in slow-wave activity.

Here, we used 256-channel high-density EEG to characterize baseline NREM oscillatory organization in children with DOA and age- and sex-matched healthy controls. We quantified spectral activity and its overnight dynamics, slow-wave amplitude and synchronization, spatial involvement and propagation, slow-wave origin and termination patterns, and Type 1 and Type 2 slow-wave properties, together with slow and fast spindle density. We specifically tested whether pediatric DOA is associated with enhanced local slow-wave synchronization but reduced large-scale recruitment, and whether this phenotype preferentially affects NREM oscillatory processes associated with widespread cortical and thalamocortical coordination.

## Methods

### Participants

Seventeen children with DOA (Amacker et al., 2026) and seventeen age- and sex-matched healthy controls (Castelnovo et al., 2023) were recruited at the Sleep Medicine Unit of the Neurocenter of Southern Switzerland as part of two studies conducted between August 2015 and February 2025. The study sample was constrained by the feasibility of acquiring overnight 256-channel hdEEG recordings in a well-characterized pediatric clinical population. This sample size is consistent with previous mechanistic hdEEG investigations of pediatric NREM slow-wave organization (Castelnovo et al., 2023; Kurth et al., 2017), as well as prior clinical EEG and hdEEG studies of NREM parasomnias in adults (i.e. Castelnovo et al., 2023; Gaudreau et al., 2000). EEG acquisition hardware, electrode net, impedance protocol, recording procedures, and preprocessing pipeline were identical across studies and remained unchanged throughout the recruitment period. Participants with DOA were recruited based on the International Classification of Sleep Disorders, Third Edition (ICSD-3) diagnostic criteria. Inclusion criteria comprised a clinical diagnosis of DOA and an age between 5-17 years. Exclusion criteria included major psychiatric or neurological disorders (e.g., epilepsy), use of psychotropic medications, and clinically significant sleep-disordered breathing, defined as an apnea-hypopnea index (AHI) >5 events/hour. Following an initial in-person or telephone screening, all patients, and when available their parents, underwent a comprehensive clinical assessment conducted by a physician board-certified in sleep medicine. Control participants were screened to exclude the presence of sleep, neurological, psychiatric, or other medical conditions potentially affecting sleep. Parents of pediatric controls underwent a structured telephone interview conducted by a pediatric sleep expert and pediatric psychiatrist (S.M.) to ensure the absence of sleep disorders, neuropsychiatric comorbidities, or relevant medical conditions in their children. Written informed consent was obtained from all participants prior to study initiation. Study procedures were reviewed and approved by the local Independent Ethics Committee “Comitato Etico Cantonale” (2015-n.2881 / 2017-n.3282), following the regulatory requirements of Switzerland and conducted in accordance with the Declaration of Helsinki.

### Procedure

All enrolled patients and controls underwent one overnight polysomnography (PSG) recording in the sleep laboratory (approximately 9:00 pm to 7:00 am). EEG was recorded using a 256-channel high-density system (Electrical Geodesics Inc., Eugene, OR), referenced to the vertex (Cz) electrode, and sampled at 500 Hz. Sleep staging was performed by a board-certified sleep medicine physician (A.C.) according to standard AASM criteria using EMBLA-RemLogic software (American Academy of Sleep Medicine, 2023). Sleep staging was conducted on 30-second epochs using six standard EEG derivations extracted from the high-density EEG system and re-referenced in a bipolar montage (F3/M2, F4/M1, C3/M2, C4/M1, O1/M2, O2/M1), synchronized with the remaining PSG channels. DOA episodes were scored by three independent raters certified in sleep medicine. For the detailed scoring procedure, see Amacker et al., 2026.

### EEG preprocessing

EEG recordings were preprocessed using the EEGLAB toolbox (version 2023.1) for MATLAB (The MathWorks, Inc., v2025b), largely following a previously validated pipeline (Michalak et al., 2026). First, the signal was downsampled to 250 Hz and high-pass filtered at 0.3 Hz using a finite impulse response (FIR) filter implemented in EEGLAB (*pop_eegfiltnew*, default settings). The filter was designed as a Hamming-windowed sinc FIR filter, with the filter order automatically determined by the software. Line noise was subsequently attenuated using the ZapLine-Plus algorithm, an adaptive approach that identifies and subtracts narrow-band noise components in the frequency domain while preserving the integrity of the broadband EEG signal (De Cheveigné, 2020; Klug & Kloosterman, 2022). To identify potential bad channels, a copy of the partially preprocessed data was re-referenced to the average of all electrodes and high-pass filtered at 1 Hz (FIR filter, *pop_eegfiltnew*). Bad channels were then detected using the findNoisyChannels function from the PREP-pipeline plugin, which applies multiple complementary criteria - including robust deviation, high-frequency noise, inter-channel correlation, and RANSAC-based spatial predictability - to identify artifactual or poorly recording electrodes (Bigdely-Shamlo et al., 2015). Channels identified as bad were excluded from subsequent processing steps and flagged for later interpolation. Residual EEG artifacts were addressed through an ICA-based cleaning procedure. To this purpose, a copy of the EEG signal was constructed retaining only NREM epochs (stages N1-N3) free of manually identified arousals, and only good channels - excluding previously identified bad channels and the recording reference - were entered into the decomposition. Prior to ICA, the data were band-pass filtered between 0.5 and 45 Hz (FIR filters, *pop_eegfiltnew*), and dimensionality was reduced to 160 principal components via PCA to reduce computational load. Independent components (ICs) were classified using the ICLabel plugin (Pion-Tonachini et al., 2019). Components for which the probability associated with the ‘brain’ label fell below an empirically determined threshold of 0.25, or for which the ratio between the brain-label probability and the highest noise-label probability was below 0.80, were rejected as artifactual. A manual override step was additionally available to correct misclassifications upon visual inspection. The ICA weights obtained from the artifact-free epochs were subsequently applied to the full, uncut continuous recording to remove artifactual components from the entire signal. Finally, bad channels were interpolated using spherical splines derived from the activity of neighboring electrodes (*eeg_interp*, ‘spherical’ method).

### Power spectral analysis

Sleep EEG power spectral density (PSD) was computed using Welch’s method, implemented via MATLAB’s *pwelch* function. Prior to any spectral estimation, EEG data (already high-pass filtered at 0.3 Hz) were low-pass filtered at 45 Hz and re-referenced to the average. Segments contaminated by arousals were excluded from the analysis. For each subject, two regions of interest (ROIs) were defined based on the GSN HydroCel 257-channel montage: a frontal ROI (22 channels) and a central ROI (15 channels). For each channel, the NREM time series was extracted and detrended, and its power spectral density was estimated using Welch’s method with a 4-second Hamming window and 50% overlap. Channel-level power spectra were then averaged across all channels within each ROI to obtain a single ROI-level power spectrum per subject, and the resulting values were subsequently converted to decibels (dB, 10·log□□) and the analysis was restricted to the 0-30 Hz frequency range. Group-level comparisons between DOA patients and controls were performed using a non-parametric cluster-based permutation test implemented in FieldTrip, with 10,000 permutations and a two-tailed cluster-corrected threshold of α = 0.05.

To characterize the spatial distribution of spectral power across the scalp, PSD was computed for five canonical frequency bands (delta: 0.5-4 Hz, theta: 4-7 Hz, alpha: 7-10 Hz, sigma: 10-16 Hz, beta: 16-30 Hz) using trapezoidal integration of the PSD (*trapz* function) within each band’s frequency range, separately for each channel and epoch. Only NREM epochs (N1, N2, N3) recorded across the whole night were retained, excluding arousal segments. Band power was then averaged across all valid NREM epochs for each channel.

### Overnight homeostatic decay

To characterize overnight changes in delta power, delta activity (0.5-4 Hz) was quantified epoch-by-epoch and channel-by-channel across the night. After average referencing and low-pass filtering (45 Hz), the continuous EEG was segmented into consecutive, non-overlapping 30-s epochs. For each epoch and each channel, PSD was estimated using Welch’s method with 4-s Hann windows and 50 % overlap within epoch; absolute delta power was then obtained by trapezoidal integration of the PSD between 0.5 and 4 Hz and expressed in decibels (10·log□□). To assess sleep homeostatic processes, we divided the whole NREM period (N1, N2 and N3 epochs, excluding those containing scored arousals) into ten sequential windows containing an equal number of epochs, according to the chronological order of NREM epochs. Dividing sleep by NREM progression rather than by clock time aligns subjects on the internal structure of their sleep, so that the early and late windows reflect comparable phases of the night across individuals despite differences in total sleep time, sleep-onset latency, NREM/REM cycling and bedtime. Within this partition, N3 epochs falling in an early NREM window (deciles 2-4) and in a late NREM window (deciles 8-10) were identified. Since the amount of N3 sleep typically decreases across the night, the number of N3 epochs available was usually larger in the early than in the late window. To avoid biases arising from unequal sampling, the number of N3 epochs was matched between the two windows within each subject. A minimum of five minutes (10 epochs) of N3 sleep in each window was required for inclusion. Mean delta power (dB) was computed separately for the early and late N3 segments, and the early-to-late difference (Δdelta = early − late) was used to quantify the overnight reduction in N3 delta power. This analysis was conducted in two ways: using absolute delta power values, to quantify the magnitude of overnight dissipation; and using across-channel z-scored values, to characterize the topographic distribution of the homeostatic effect independently of interindividual differences in overall power magnitude.

### Slow wave detection

Following preprocessing, the EEG signal was band-pass filtered between 0.5 and 45 Hz (FIR filters, *pop_eegfiltnew*) and re-referenced to the average of the two mastoid electrodes. Slow wave detection was performed using a custom MATLAB algorithm (Bernardi et al., 2018; Mensen et al., 2016). Prior to detection, a baseline correction was applied by subtracting the channel-wise median after removing extreme artifacts (|negative amplitude| > 400 µV). A slow trend was removed by subtracting the median computed over 10-second windows. A candidate event was retained as a slow wave if its half-wave duration fell between 250 ms and 1000 ms (full -wave period 0.5 - 2.0 s or 0.5 - 2.0 Hz frequency range). In line with our previous works, no amplitude threshold was used (Bergamo et al., 2026; Bernardi et al., 2018; Castelnovo et al., 2023; Spiess et al., 2018). For each detected wave, the following features were identified: the negative zero-crossing, the positive zero-crossing, the maximal negative peak, the maximal positive peak, and the maximum downward and upward slopes. To distinguish the local strength of slow-wave synchronization from their large-scale spatial organization, slow waves were characterized using complementary spatial-envelope and channel-wise approaches. In the spatial-envelope approach, a spatial envelope signal was derived by sorting all channels by instantaneous amplitude at each time point and averaging the four most negative channels, excluding the single most negative, which was treated as potentially artifactual (Bergamo et al., 2024). In the channel-wise approach, detection was applied independently to each channel (Riedner et al., 2007). In both cases, detected slow waves were subsequently restricted to NREM sleep epochs (stages N1-N2-N3) free of manually scored arousals.

### Classification of slow waves by synchronization profile

For each detected slow wave, a synchronization score was computed to quantify both the number of cortical units contributing to the wave and the rapidity of their synchronization (Bernardi et al., 2018; Castelnovo et al., 2023; Siclari et al., 2018). For each wave, the mean EEG amplitude across all scalp electrodes (excluding mastoids and peripheral on the neck and cheeks) was computed in a ±20 ms window centered on the negative peak. The involvement was defined as the fraction of channels showing a negative deflection exceeding 5 µV at the time of the negative peak. The synchronization score was then calculated as the product of involvement and the mean maximum downward and upward slopes, providing a single scalar measure combining spatial extent and wave steepness. To distinguish events with high versus intermediate synchronization profiles, waves at or above the participant-specific 90th percentile of the synchronization-score distribution were classified as Type 1, whereas those between the 45th and 55th percentiles were classified as Type 2 (Castelnovo et al., 2023). Participant-specific thresholds were used to account for interindividual differences in the absolute distribution of synchronization scores.

### Slow wave propagation

To characterize the spatiotemporal propagation of slow waves across the scalp, a delay map was computed for each detected event following a validated approach (Bernardi et al., 2018). For each wave, the EEG signal was extracted in a symmetric window of ±500 ms around the negative peak identified from the reference envelope detection. The local negative peak latency was then determined for each electrode within a ±150 ms window around the reference peak, using peak detection on the smoothed signal. A likeness constraint was applied to exclude channels whose waveform deviated substantially from a prototype slow wave, defined as the wave with the largest negative amplitude across participating channels (Menicucci et al., 2009; Mensen et al., 2016). Likeness was quantified via cross-correlation between each channel’s signal and the prototype within a symmetric ±150 ms window; only channels exceeding the 25th percentile of the cross-correlation distribution were retained. To exclude propagation gaps potentially reflecting local artifacts, a spatiotemporal clustering procedure was applied: two electrodes were considered part of the same propagation cluster if their negative peak latencies differed by less than ±12 ms and they were spatial neighbors according to a pre-computed channel adjacency structure (Avvenuti et al., 2020). The cluster containing the channels with the earliest negative peak was retained while all the other channels were set to NaN. The minimum delay across participating channels was then set to zero to obtain the final delay map. Origin and termination maps were defined probabilistically: a channel was classified as an origin if its delay was ≤4 ms, and as a termination if its delay was ≥20 ms. Origin probability of a given electrode was computed as the proportion of all detected slow waves originating at that channel. Origin probability therefore represents the relative spatial distribution of slow-wave origins across the scalp. These measures were used to characterize the spatial extent and temporal recruitment of scalp regions during individual slow waves, rather than functional connectivity between cortical regions.

### Sleep spindle detection

Sleep spindles were detected using a continuous wavelet transform (CWT)-based algorithm, targeting the 10-16 Hz frequency range, applied independently to each EEG channel (Bergamo et al., 2026; Mensen et al., 2018). Events were retained if their duration was between 300 ms and 3 seconds. A spectral quality criterion was additionally applied: for each candidate spindle, a power spectral density estimate (Welch’s method) was computed on the spindle segment, and the event was accepted only if the mean power exceeded that of the neighboring frequency bands (8-10 Hz and 16-18 Hz) by a factor of 2. For each retained spindle, the following features were extracted: start and end sample, wavelet peak time and peak frequency. Only events identified in N2 or N3 sleep were retained, excluding arousals. Spindles were further classified according to their peak frequency as slow (<13 Hz) or fast (≥13 Hz) spindles and were analyzed separately within lighter (N1+N2) and deeper (N3) sleep stages.

### Statistics

Statistical analyses were two-sided, with significance set at p < 0.05. Group differences in conventional sleep parameters were assessed using independent-samples t-tests or Mann-Whitney U tests, according to normality assessed with the Shapiro-Wilk test. Multiple comparisons across sleep parameters were controlled using the Benjamini-Hochberg false discovery rate (FDR; Benjamini & Hochberg, 1995), with significance defined as q < 0.05. For topographic EEG measures, group effects were estimated using robust linear regression models including age and sex as covariates. Spatial multiple comparisons were controlled using cluster-based permutation testing across electrodes. Electrodes exceeding an uncorrected two-sided threshold of p < 0.05 were grouped according to spatial adjacency, with cluster mass defined as the sum of the corresponding test statistics. Statistical significance was determined against a null distribution obtained from 10,000 permutations of group labels, with cluster-corrected p < 0.05 considered significant (two-tailed, an approach already used in Bergamo et al., 2026). For non-topographic slow-wave and spindle measures, group differences were assessed using robust linear regression models including age and sex as covariates. Benjamini-Hochberg FDR correction was applied within each sleep stage (e.g., N3 and N2+N1), with q < 0.05 considered significant.

## Results

### Sleep architecture

The DOA group (n = 17, 8F/9M) and the control group (n = 17, 7F/10M) were comparable for age (DOA: 11.3 ± 2.9 years; controls: 10.4 ± 1.7 years; t = 1.00, p = 0.325) and sex distribution (χ² = 0.51, p = 0.472). The two groups did not differ in apnea-hypopnea index (DOA: 0.9 ± 0.8; controls: 0.7 ± 0.6; U = 310.5, p = 0.666), confirming the absence of clinically relevant sleep-disordered breathing in both samples. Patients with DOA exhibited 2.9 ± 2.5 overnight episodes. Conventional sleep architecture was largely comparable between groups (Table 1). Although children with DOA showed nominally shorter total sleep time and sleep-onset latency, longer REM latency, and shorter N3 duration, none of these differences survived FDR correction (all q ≥ 0.179). No group differences were observed in sleep efficiency, relative sleep-stage composition, arousal index, or NREM awakening indices (all q ≥ 0.537).

**Table 1.** Demographic, clinical, and polysomnographic characteristics of children with disorders of arousal (DOA) and healthy controls. Values are reported as mean ± standard deviation (M ± SD), unless otherwise indicated. Group differences were assessed using independent-samples *t*-tests, Mann-Whitney *U* tests, or chi-square (χ²) tests, as appropriate. *P* values surviving false discovery rate (FDR) correction are reported in the last column. Significant uncorrected comparisons are indicated with an asterisk (p < 0.05). Abbreviations: AHI, apnea-hypopnea index; TST, total sleep time; WASO, wake after sleep onset; SOL, sleep onset latency; REML, REM sleep latency; SE, sleep efficiency; AI, arousal index; AWI, awakening index; NREM, non-rapid eye movement sleep; REM, rapid eye movement sleep.

| | DOA (M ± SD) | CONTROLS (M ± SD) | test | p-value | FDR corrected p-value |
| --- | --- | --- | --- | --- | --- |
| Age | 11.3 ± 2.9 | 10.4 ± 1.7 | $t = 1$ | 0.325 | |
| Sex | 8F/9M | 7F/10M | $\chi^2 = 0.51$ | 0.472 | |
| Episodes | 2.9 ± 2.5 | n.a. |  |  |  |
| AHI | 0.9 ± 0.8 | 0.7 ± 0.6 | U = 310.5 | 0.666 |  |
| TST (min) | 409.5 ± 34.9 | 431.4 ± 53.3 | U = 85 | 0.042 * | 0.179 |
| WASO (min) | 35.3 ± 30.5 | 44.3 ± 40.3 | U = 139.5 | 0.877 | 0.982 |
| SOL (min) | 18.8 ± 20.3 | 36.0 ± 24.4 | U = 77.50 | 0.022 * | 0.179 |
| REML (min) | 116.5 ± 51.6 | 80.6 ± 24.2 | t = 2.59 | 0.014 * | 0.179 |
| SE (%) | 88.5 ± 6.2 | 84.5 ± 9.1 | U = 186 | 0.158 | 0.537 |
| N1 (%) | 6.4 ± 2.4 | 6.2 ± 2.9 | U = 160.00 | 0.605 | 0.859 |
| N2 (%) | 39.3 ± 5.8 | 37.5 ± 4.6 | t = 1.00 | 0.323 | 0.784 |
| N3 (%) | 32.3 ± 4.6 | 34.0 ± 3.9 | t = -1.21 | 0.237 | 0.671 |
| REM (%) | 22.0 ± 3.8 | 22.3 ± 5.1 | t = -0.19 | 0.854 | 0.982 |
| N1 (min) | 26.3 ± 10.1 | 26.2 ± 11.4 | t = 0.01 | 0.994 | 0.994 |
| N2 (min) | 160.7 ± 25.2 | 161.5 ± 25.8 | t = -0.10 | 0.924 | 0.982 |
| N3 (min) | 131.7 ± 18.7 | 145.8 ± 18.9 | t = -2.18 | 0.037 * | 0.179 |
| REM (min) | 90.8 ± 20.7 | 97.9 ± 30.0 | t = -0.80 | 0.430 | 0.784 |
| AI (#/h) | 12.3 ± 3.0 | 12.8 ± 3.0 | t = -0.45 | 0.655 | 0.859 |
| AWI NREM (#/h) | 2.7 ± 0.9 | 2.5 ± 0.8 | t = 0.79 | 0.436 | 0.784 |
| AWI N1N2 (#/h) | 2.2 ± 0.8 | 2.0 ± 0.7 | t = 0.75 | 0.461 | 0.784 |
| AWI N3 (#/h) | 0.5 ± 0.3 | 0.4 ± 0.3 | t = 0.45 | 0.657 | 0.859 |

### NREM spectral power is globally increased in pediatric DOA

Spectral analysis of whole-night NREM EEG revealed a broad increase in low-frequency power in children with DOA relative to controls (Figure 1a). Power spectra extracted from both the central and frontal ROIs showed significantly higher power in DOA across the delta, theta, and alpha bands (∼0-10 Hz) (Central ROI: cluster-mass = 125.26, cluster-corrected *p* = 0.003; Frontal ROI: cluster mass = 128.60, cluster-corrected p = 0.002). A second, narrower significant cluster was also identified in the sigma/low-beta range (∼13-21 Hz) (Central ROI: cluster-mass = 77.83, cluster-corrected *p* = .013; Frontal ROI: cluster mass = 92.49, cluster-corrected p = 0.007). Topographical analysis of integrated power for each frequency band confirmed this pattern (Figure 1b): DOA patients showed diffusely higher delta, theta and alpha power across nearly the entire scalp, with cluster mass-corrected differences in frontal, central and parietal derivations (delta: cluster-mass = 291.84, cluster size = 108 channels, cluster-corrected *p* = 0.014; theta: cluster-mass = 568.31, cluster size = 171 channels, cluster-corrected *p* = 0.002; alpha: cluster-mass = 514.62, cluster size 172 channels, cluster-corrected *p* = 0.007). By contrast, sigma-band differences did not survive cluster correction (cluster-mass = 7.32, cluster size = 3 channels, cluster-corrected *p* = 0.294), whereas beta-band effects remained restricted to a smaller fronto-central cluster (cluster-mass = 91.09, cluster size = 37 channels, cluster-corrected *p* = 0.031). After across-channel z-score normalization, no significant group differences remained in any frequency band (all cluster-corrected p > 0.05), indicating that these effects reflected differences in overall NREM power magnitude rather than its relative scalp distribution.

**Figure 1.**
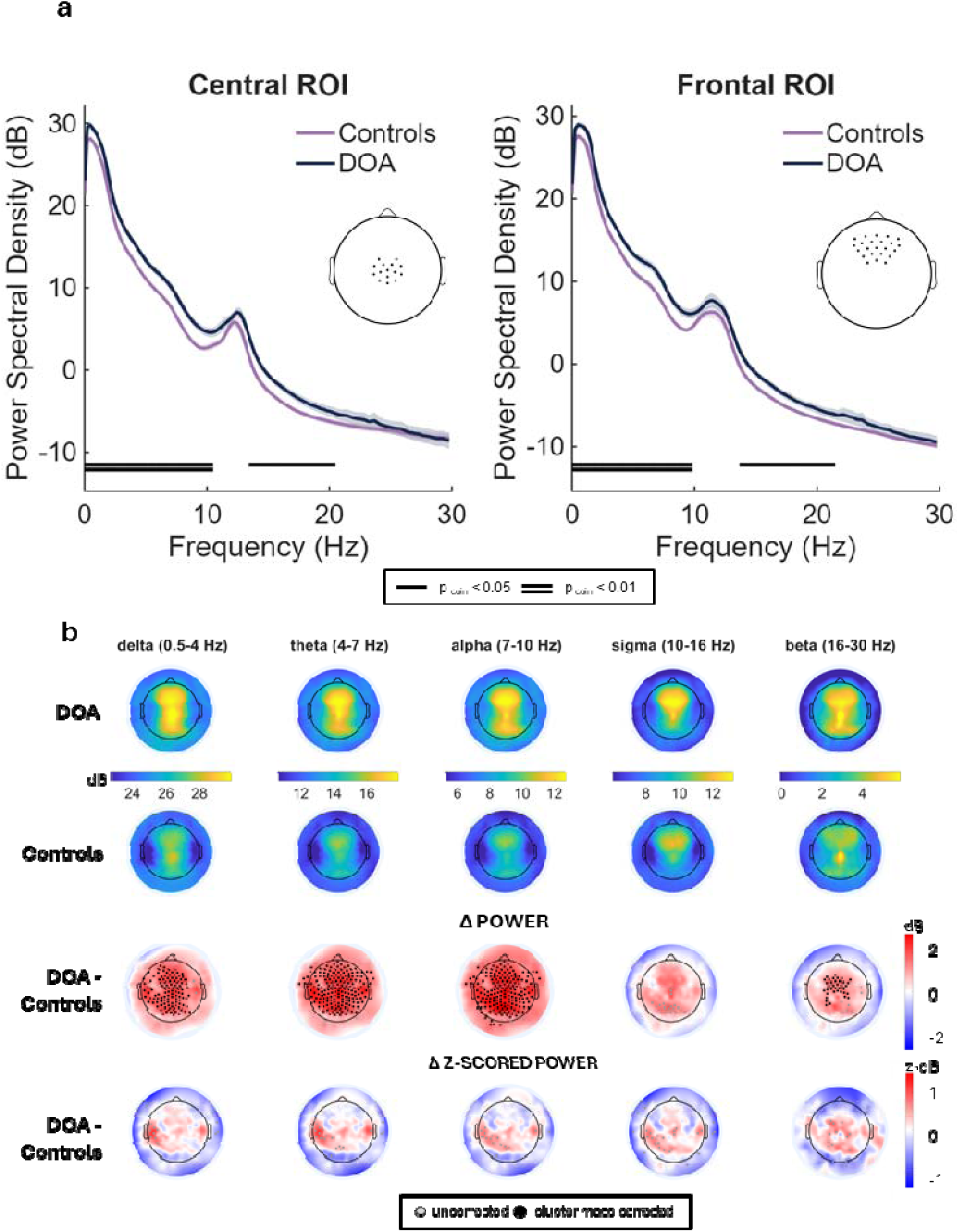
Whole-night NREM spectral power in children with DOA vs controls. **a)** Power spectra averaged across a central and a frontal ROI. Children with DOA (blue) exhibited significantly higher spectral power than controls (purple), with two significant clusters in the ∼0-10 Hz and ∼13-21 Hz frequency ranges (cluster-based permutation test, cluster-corrected *p* < 0.05; black bars indicate significant frequency clusters). Shaded areas represent mean ± SE. Insets illustrate the electrode locations included in each ROI. **b)** Topographic distribution of spectral power for delta (0.5-4 Hz), theta (4-7 Hz), alpha (7-10 Hz), sigma (10-16 Hz) and beta (16-30 Hz) bands. Upper panels show absolute band power for each group. Middle panels display group differences (Δ spectral power, DOA - controls). The lower panels show the corresponding z-scored group differences (Δ z-scored spectral power, DOA - controls). Black dots indicate channels belonging to significant clusters after cluster-level correction, whereas gray dots indicate uncorrected significance.

### Overnight decline in delta power is preserved in DOA

Delta power decreased significantly from early to late N3 sleep across nearly the entire scalp in both groups (both cluster-corrected p < 0.001; Figure 2). Children with DOA showed higher absolute delta power during early N3 over a centro-parietal cluster (cluster-corrected p = 0.021), whereas no significant group difference was observed during late N3. Although the magnitude of overnight SWA decline (Δ early-late) was higher in DOA patients than in controls, this effect did not survive correction for multiple comparisons (all corrected *p* > 0.05). Similarly, after across-channel z-score normalization, no between-group differences were detected in early, late, or early-to-late delta-power topographies (all cluster-corrected p > 0.05). Additionally, controls showed a significant overnight shift toward relatively greater centro-parietal SWA (cluster mass = 80.57, cluster size = 24, *p* = 0.002), whereas children with DOA showed a significant fronto-central cluster in the z-scored SWA change (cluster mass = 67.41, cluster size = 22, *p* = 0.008), reflecting a relative redistribution of SWA over anterior regions. However, direct comparisons of the early, late, and Δ z-scored topographies revealed no significant group differences (all cluster-corrected *p* > 0.05). Thus, although the within-group patterns of overnight SWA redistribution differed topographically, these differences were not statistically supported at the between-group level.

**Figure 2.**
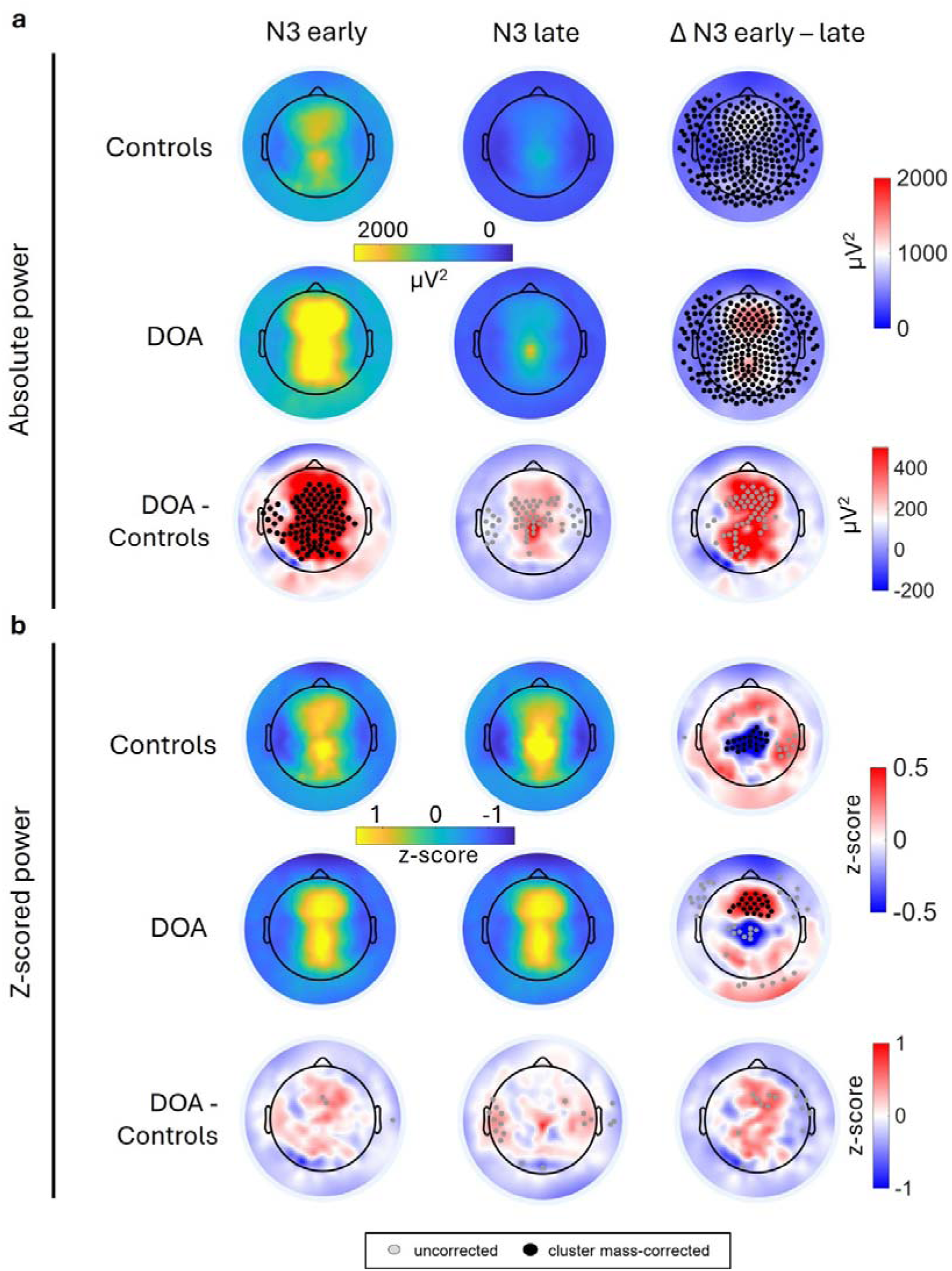
Overnight homeostatic decay and topographic redistribution of slow-wave activity in DOA and controls. Topographic maps of delta power during early (N3 seg. 2-4) and late (N3 seg. 8-10) sleep and their difference (Δ), shown as absolute (**a**) and z-scored (**b**) power for controls, DOA, and their contrast (DOA - Controls). Black filled dots indicate electrodes within significant clusters (cluster-corrected *p* < 0.05); grey dots show uncorrected channels (*p* < 0.05 uncorrected).

### Enhanced local slow-wave expression coexists with reduced large-scale recruitment in DOA

Channel-wise analyses revealed a spatial reorganization of slow-wave activity in children with DOA (Figure 3a). Slow-wave origin probability was redistributed toward the central vertex region, with lower relative origin probability across most of the remaining scalp. This pattern was significant across all NREM sleep (cluster mass = 551.92, cluster size = 177, cluster-corrected *p* < 0.001), as well as during N1+N2 (cluster mass = 545.71, cluster size = 171, cluster-corrected *p* < 0.001) and N3 sleep (cluster mass = 547.61, cluster size = 182, cluster-corrected *p* < 0.001). In parallel, slow-wave amplitude was higher in DOA over central and centro-parietal regions across all NREM sleep (cluster mass = 193.78, cluster size = 78, cluster-corrected *p* = 0.025), with similar differences during N1+N2 (cluster mass = 280.33, cluster size = 105, cluster-corrected *p* = 0.009) and N3 sleep (cluster mass = 199.24, cluster size = 80, cluster-corrected *p* = 0.024). Slow-wave density was also higher in DOA, reaching cluster-corrected significance during N3 over central and posterior regions (cluster mass = 441.81, cluster size = 175, cluster-corrected *p* = 0.011), whereas involvement showed no consistent group differences.

**Figure 3.**
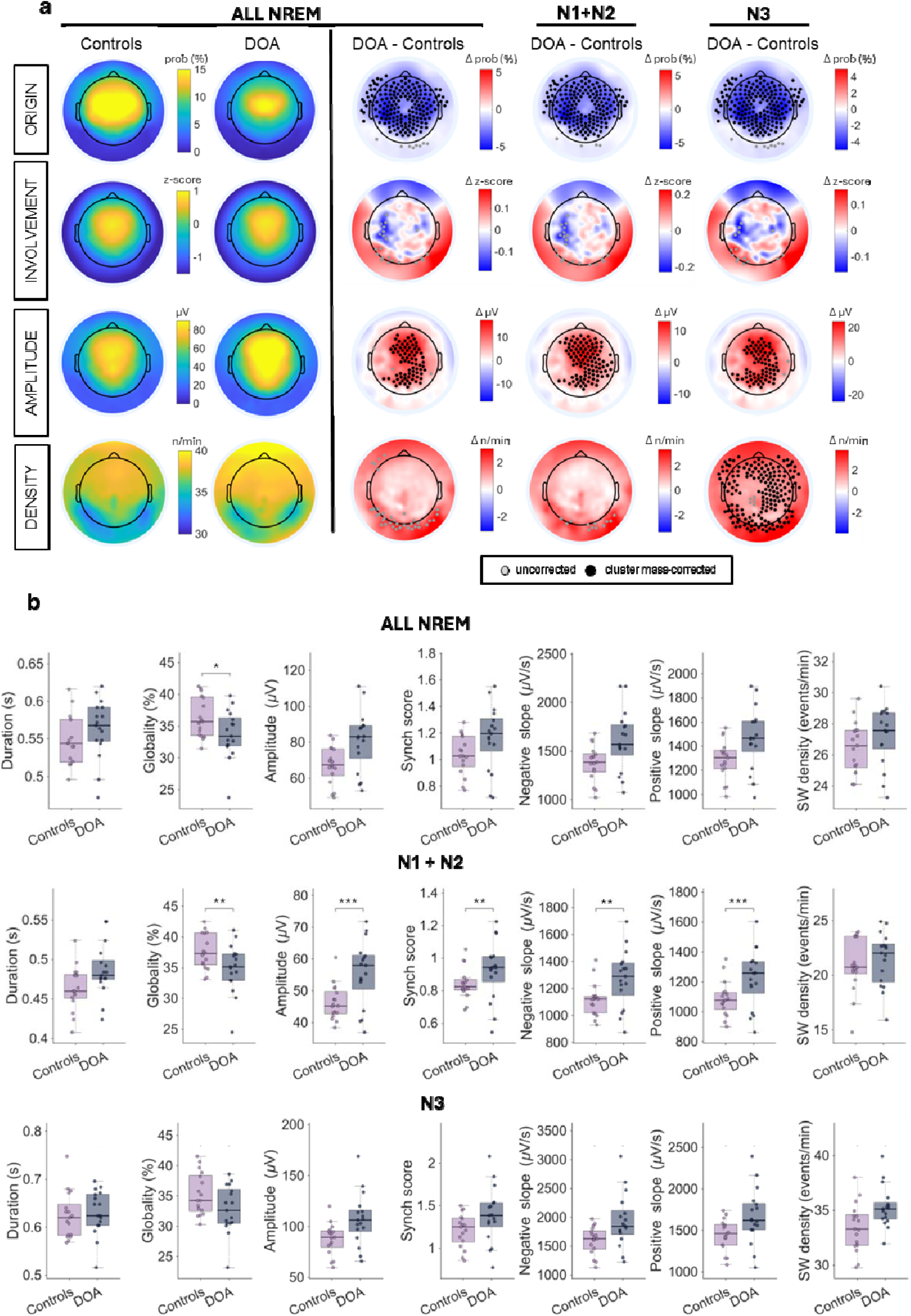
Slow wave characteristics in DOA patients vs Controls. **a)** Topographic maps of four slow-wave parameters - origin, involvement, amplitude, and density. The first two columns show group-average values for controls and DOA across all NREM sleep. The remaining columns show between-group differences (DOA-Controls) for all NREM, for N1+N2, and for N3 sleep. Black filled dots indicate electrodes within significant clusters (cluster-corrected *p* < 0.05); grey dots show uncorrected channels (*p* < 0.05 uncorrected). **b)** Box plots of slow-wave parameters as detected on the signal envelope - duration, globality, amplitude, synchronization score (synch score), negative slope, positive slope, and density-for controls versus DOA, shown separately for ALL NREM, N1+N2, and N3. Each dot represents an individual participant. Boxes show the median and interquartile range; whiskers extend to the most extreme values within 1.5 × IQR. *q < 0.05; **q < 0.01; ***q < 0.001.

At the level of individual spatial-envelope events, a dissociation emerged between the strength of slow-wave synchronization and its spatial extent (Figure 3b). This pattern was most evident during N1+N2 sleep, when slow waves in DOA showed greater amplitude (β = 11.46, *t*(30) = 4.22, *q* < 0.001), higher synchronization scores (β = 0.13, *t*(30) = 2.95, *q* = 0.008), and steeper negative (β = 178, *t*(30) = 3.01, *q* = 0.008) and positive slopes (β = 199.26, *t*(30) = 4.12, *q* < 0.001), while exhibiting lower globality (β = −3.21, *t*(30) = −3.03, *q* = 0.008). Across all NREM sleep, reduced globality remained significant (β = −3.26, *t*(30) = −2.90, *q* = 0.048), whereas no envelope-derived slow-wave measure differed significantly between groups during N3 after correction for multiple comparisons. Thus, the envelope analysis revealed that greater slow-wave synchronization in DOA was not accompanied by greater spatial recruitment and was most evident during lighter NREM sleep.

We next examined whether this phenotype differed according to the slow-wave synchronization profile. The overall distribution of synchronization scores in DOA showed a lower median peak but a heavier right tail (Figure 4a). When waves were classified using participant-specific synchronization thresholds, between-group differences were predominantly observed among Type 1 waves (Figure 4b). In DOA, Type 1 waves showed greater amplitude (β = 38.11, *t*(30) = 2.80, *q* = 0.009) and steeper positive (β = 518.54, *t*(30) = 3.72, *q* = 0.004) and negative slopes (β = 795.24, *t*(30) = 2.98, *q* = 0.009), together with lower globality (β = −5.29, *t*(30) = −2.79, *q* = 0.009). By contrast, differences in amplitude and globality among Type 2 waves were observed only at the uncorrected level and did not survive correction for multiple comparisons. Slow-wave origin probability was redistributed toward central regions in both Type 1 and Type 2 waves (Type 1: cluster mass = 336.04, cluster size = 124, cluster-corrected *p* = 0.002; Type 2: cluster mass = 462.51, cluster size = 153, cluster-corrected *p* < 0.001). As a sensitivity analysis, we repeated slow-wave analyses within the first hour of NREM sleep, when sleep pressure is highest and DOA episodes preferentially occur. The overall pattern was preserved, with additional differences emerging for envelope-derived N3 measures and Type 2 waves (Supplementary Fig. S1).

**Figure 4.**
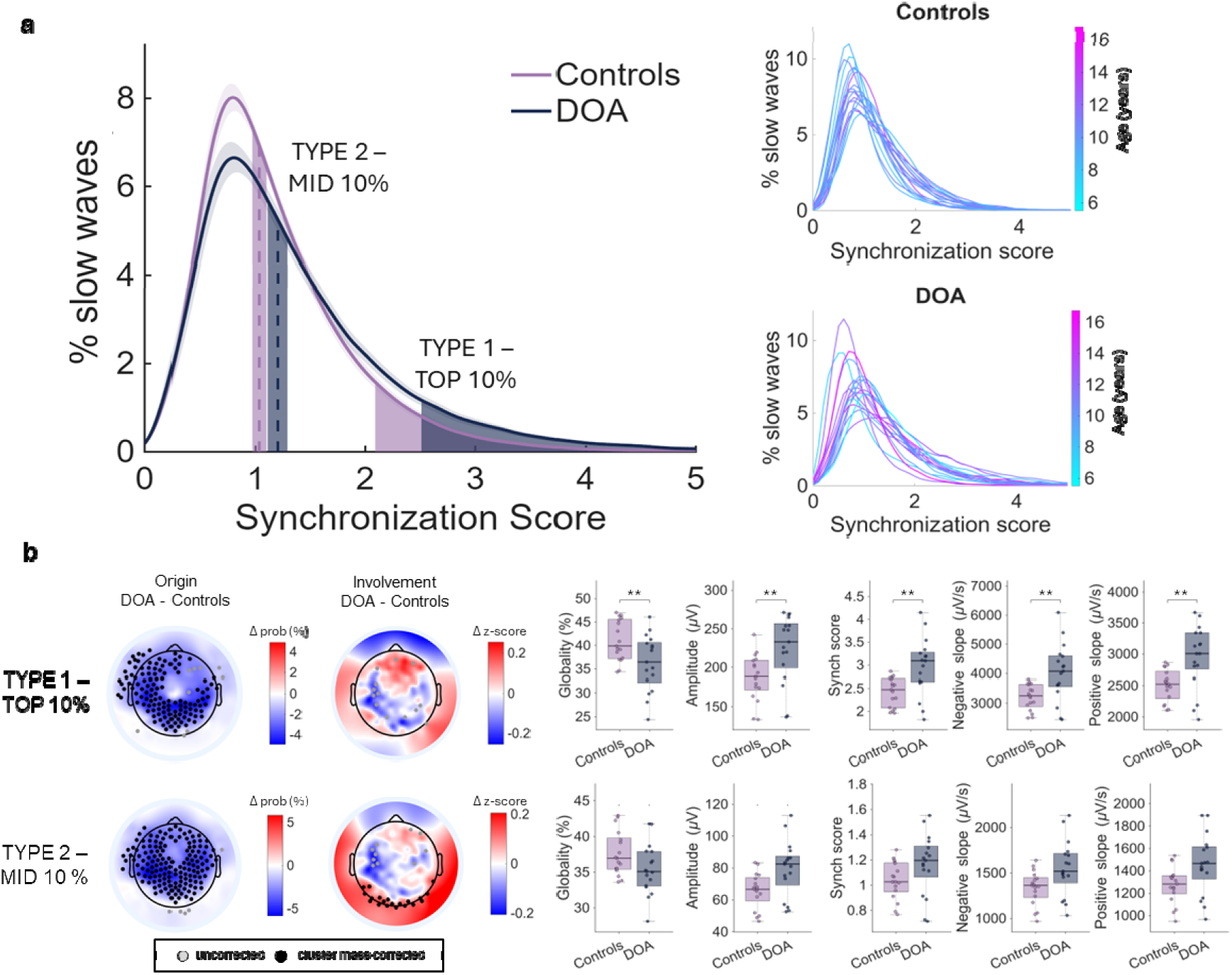
Slow waves sub-types. **a)** Slow wave synchronization score distribution. Left: Group distribution of synchronization scores (mean ± SD). Dashed vertical lines indicate group-specific median values. Shaded areas highlight percentile-based thresholds used to define the mid-range (45-55th percentile - MID 10%) and the top 10% of synchronization scores (>90 th percentile - TOP 10%). Right: individual distributions of synchronization scores for controls (top) and DOA (bottom), color-coded by participant age (years). **b)** Between-group differences (DOA-Controls) separated by slow-wave type. TYPE 1 (top) and TYPE 2 (bottom) are shown with topographic maps of origin and involvement (uncorrected and cluster mass-corrected electrodes) and box-plots of globality, amplitude, synchronization score, and negative and positive slopes. Black dots indicate electrodes within significant clusters (cluster-corrected *p* < 0.05); grey dots show uncorrected channels (*p* < 0.05 uncorrected). *q < 0.05; **q < 0.01; ***q < 0.001.

### Centro-posterior slow spindle density is reduced during N3 in DOA

The density of slow spindles, which represent the majority of sleep spindles in both groups (Supplementary Table S1), displayed the expected anterocentral topographic maximum, whereas the density distribution of fast spindles was more spatially circumscribed and centroparietally distributed. Spindle density was lower in DOA across much of the scalp, but between-group differences survived cluster correction only during N3 sleep over centro-posterior regions (Figure 5). This effect was observed for overall spindle density (cluster mass = 213.91, cluster-corrected p = 0.038) and slow spindles (<13 Hz; cluster mass = 222.28, p = 0.037). By contrast, fast spindle density did not differ significantly between groups in any stage (all p > 0.05).

**Figure 5.**
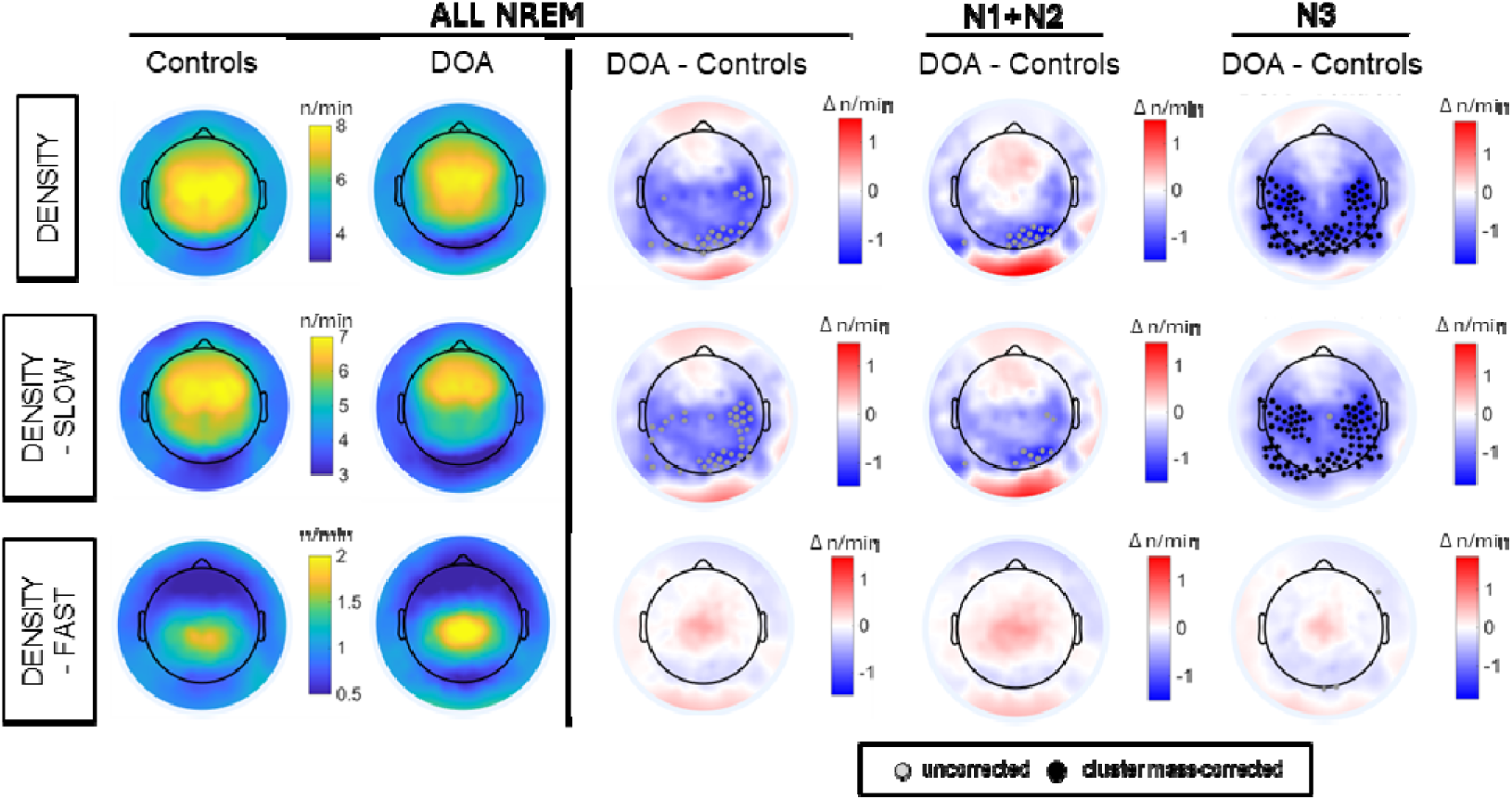
Topographic distribution of spindle density across all NREM sleep and separately for sleep stages in DOA patients and controls. Topographic maps of spindle density during all NREM sleep, N1+N2, and N3, shown as overall (DENSITY), slow (DENSITY - SLOW), and fast (DENSITY - FAST) spindles. Absolute density (spindles/min) is shown for controls and DOA (left), alongside the DOA - Controls contrast (Δ) for each sleep stage. Black dots indicate electrodes within significant clusters (cluster-corrected *p* < 0.05); grey dots show uncorrected channels (*p* < 0.05 uncorrected).

## Discussion

The present study reveals a dissociation between the local strength and large-scale spatial organization of NREM slow waves in children with DOA. Despite largely preserved conventional sleep architecture, children with DOA showed globally increased NREM oscillatory power and slow waves that were locally larger, steeper and more synchronized, yet less globally expressed. This phenotype was most prominent among Type 1 slow waves and was accompanied by reduced centro-posterior slow spindle density during N3 sleep. Together, these findings extend previous evidence that the local strength of slow-wave synchronization and its large-scale spatial expression represent dissociable dimensions of NREM activity (Castelnovo et al., 2023), showing that this local-to-global balance is selectively altered in pediatric DOA.

Several observations in our data converge on this interpretation. Slow-wave amplitude and slope are commonly considered indices of the strength and synchrony of local cortical activity, whereas spatial propagation depends on the recruitment of progressively distant cortical areas and is constrained by long-range structural connectivity (Esser et al., 2007; Riedner et al., 2007; Vyazovskiy et al., 2007). In DOA, increased spectral power and larger central and centro-parietal slow waves coexisted with reduced globality of individual waves, while slow-wave origins were more spatially restricted over central regions-a pattern also emerging in our preliminary unpublished observations from younger children. Notably, despite higher delta power during early N3, its overnight decline was largely preserved, suggesting no major alteration of homeostatic dissipation while leaving open the possibility that differences in homeostatic sleep regulation may contribute to DOA vulnerability. These microstructural abnormalities were also largely undetected by conventional polysomnography, with no significant group differences in sleep architecture or conventional arousal and awakening indices after correction for multiple comparisons. Together, these findings point to an alteration not simply in the amount or intensity of NREM sleep, but in how locally synchronized activity is organized across distributed cortical areas. A developmental perspective may be particularly relevant to this phenotype. Slow-wave activity undergoes pronounced changes throughout childhood and adolescence, in parallel with major changes in cortical anatomy and connectivity (Buchmann et al., 2011; Kurth et al., 2010, 2017). Compared with adults, children show larger and steeper but less spatially widespread slow waves, while propagation distance increases across development and has been associated with myelin content in long-range white-matter pathways (Castelnovo et al., 2023; Kurth et al., 2017; Schoch et al., 2018). Pediatric DOA may therefore reflect an altered balance between processes supporting strong local synchronization and those enabling distributed recruitment during NREM sleep. Within the broader developmental trajectory, DOA may represent an extreme neurophysiological profile characterized by a different balance between local and distributed NREM synchronization. This framework could also help contextualize the higher pediatric prevalence (Petit et al., 2015; Stallman & Kohler, 2016), and the apparently divergent findings reported across age groups, with increased slow-wave power in the present pediatric sample but reduced or altered SWA dynamics and greater NREM fragmentation reported in adults with DOA (Carpentier et al., 2020; Espa et al., 2000; Gaudreau et al., 2000; Guilleminault et al., 2001). Whether this physiological organization changes with maturation, and whether its persistence is associated with DOA in adulthood, will require longitudinal investigation.

The preferential involvement of Type 1 slow waves further supports the role of highly synchronized, widespread events to the observed phenotype. Type 1 waves are large, steep and spatially widespread slow waves associated with a bottom-up, subcortico-cortical, arousal-system-dependent synchronization process, whereas Type 2 waves were proposed to rely more strongly on horizontal cortico-cortical connections (Bernardi et al., 2018; Siclari et al., 2014). Simultaneous EEG-fMRI has further associated highly synchronized slow waves with greater thalamic involvement and markers of higher arousal and autonomic activation (Bergamo et al., 2026), while their spatial organization shows particularly pronounced developmental changes (Castelnovo et al., 2023). It is to be noted that the clearest abnormalities in DOA occurred precisely among waves normally characterized by efficient, widespread cortical recruitment: Type 1 waves were larger and steeper, but less globally expressed. The accompanying reduction in centro-posterior slow spindle density during N3, consistent with previous reports of reduced N3 spindle density in adult DOA (Carpentier et al., 2020; Gaudreau et al., 2000) provides convergent, although indirect, evidence that the phenotype is not restricted to cortical slow-wave morphology. Because spindle generation depends on coordinated thalamocortical and cortical activity, the joint alteration of slow-wave spatial organization and spindle expression may reflect altered interactions across distributed NREM oscillatory networks.

How might this altered spatial coordination during baseline NREM sleep relate to the defining phenomenon of DOA? Neuroimaging and intracranial recordings during DOA episodes have shown that wake-like activation of motor and cingulate regions can coexist with persistent sleep-like activity in associative cortical areas (Bassetti et al., 2000; Terzaghi et al., 2009, 2012), providing direct evidence for regional state dissociation during behavioral episodes and even during physiological transitions (Nobili et al., 2012). The present findings raise the possibility that such episode-related dissociation occurs against a background in which the coordination of local NREM states across distributed regions is already altered during inter-episode sleep. In this framework, vulnerability to incomplete arousal would relate not simply to the depth of local sleep, but also to the ability of distributed brain regions to coordinate changes in their physiological state. Neuromodulatory arousal systems represent one possible mechanism through which such coordination could be regulated. In particular, locus-coeruleus noradrenergic activity modulates thalamic activity, spindle clustering, autonomic activity and sensory arousability during NREM sleep (Osorio-Forero et al., 2021), and asynchronous recruitment of functionally distinct noradrenergic modules has been proposed as a potential contributor to spatially dissociated arousals in DOA (Bellesi & Castelnovo, 2026). The present data do not directly test this mechanism, but provide a physiological framework within which such hypotheses can be examined.

Some limitations should be considered. The sample size is necessarily constrained by the challenges of acquiring 256-channel hdEEG recordings in a well-characterized pediatric clinical population. Although comparable to previous mechanistic pediatric hdEEG studies, it limits the examination of clinical heterogeneity, including DOA subtype and episode frequency. The cross-sectional design precludes inference about developmental trajectories, and pubertal maturation was not directly assessed. Moreover, the developmental interpretation is based on previous evidence linking slow-wave properties and propagation to cortical and white-matter maturation rather than on structural measurements in the present participants. We therefore lack direct measures of structural or functional connectivity, as well as of the subcortical and thalamocortical mechanisms potentially underlying the observed slow-wave and spindle abnormalities. Longitudinal multimodal studies combining hdEEG with structural imaging and measures of subcortical arousal systems will be important to test these hypotheses directly.

In conclusion, pediatric DOA is characterized by a dissociation between enhanced local slow-wave synchronization and reduced large-scale spatial recruitment during NREM sleep, most prominently involving highly synchronized Type 1 waves. These findings suggest that vulnerability to incomplete arousals during childhood may emerge from an altered balance between local cortical synchronization and distributed NREM coordination during brain development. More broadly, they identify local synchronization and large-scale recruitment as dissociable dimensions of human NREM sleep whose developmental balance may contribute to the stability of sleep-wake transitions. Whether persistence of this physiological phenotype predicts DOA into adulthood remains an important question for longitudinal studies.

## Data Availability

All data produced in the present study are available upon reasonable request to the authors

## Acknowledgments

We thank the children and families who participated in this study, and the technical staff of the Sleep Medicine Unit, Neurocenter of Southern Switzerland, for their assistance with data acquisition.

## Data Availability statement

The data that support the findings of this study are available from the corresponding author upon reasonable request.

## Code Availability Statement

The codes used to preprocess and analyze the data are available upon request.

## CRediT author statement

- Conceptualization: AC, DB
- Methodology: DB, AC, GB
- Software: DB
- Formal analysis: DB
- Investigation: AC, MP
- Resources: AC, MM, SG
- Data curation: DB, AC, GL (supporting)
- Visualization: DB
- Supervision: AC
- Funding acquisition: AC, DB
- Writing - original draft: DB, AC
- Writing - review & editing: all authors

