## Supplementary Material for "Enhanced local synchronization with impaired large-scale recruitment of NREM slow waves in pediatric disorders of arousal"

| **Sleep Variables** | **DOA** | | **CONTROLS** | |
| --- | --- | --- | --- | --- |
|  | **N (event per minute)** | **SD** | **N (event per minute)** | **SD** |
| **Spindles** |  |  |  |  |
| N2 | 1224.8 (7.83 spd/min) | ± 413.3 | 1199.4 (7.61 spd/min) | ± 464.2 |
| N3 | 781.1 (6.03 spd/min) | ± 289.6 | 908.1 (6.29 spd/min) | ± 457.9 |
| NREM | 2005.9 (6.51 spd/min) | ± 612.1 | 2107.4 (6.53 spd/min) | ± 853.9 |
| **Slow spindles** |  |  |  |  |
| N2 | 1133.9 (7.25 spd/min) | ± 406.5 | 1110.4 (7.04 spd/min) | ± 437.1 |
| N3 | 724.1 (5.60 spd/min) | ± 286.7 | 839.9 (5.82 spd/min) | ± 432.7 |
| NREM | 1858.1 (6.04 spd/min) | ± 609.9 | 1950.3 (6.04 spd/min) | ± 796.1 |
| **Fast spindles** |  |  |  |  |
| N2 | 90.9 (0.58 spd/min) | ± 64.7 | 89.0 (0.57 spd/min) | ± 61.8 |
| N3 | 57.0 (0.44 spd/min) | ± 44.2 | 68.1 (0.47 spd/min) | ± 59.0 |
| NREM | 147.9 (0.48 spd/min) | ± 104.9 | 157.1 (0.48 spd/min) | ± 117.6 |
| **Slow waves** |  |  |  |  |
| N1 | 418.9 (19.17 SW/min) | ± 177.6 | 418.4 (19.14 SW/min) | ± 199.1 |
| N2 | 3419.9 (21.83 SW/min) | ± 677.4 | 3322.0 (21.13 SW/min) | ± 642.0 |
| N3 | 4573.2 (35.28 SW/min) | ± 654.8 | 4811.8 (33.44 SW/min) | ± 681.0 |
| NREM | 8412.1 (27.26 SW/min) | ± 875.6 | 8552.2 (26.48 SW/min) | ± 953.8 |
| **Type 1 slow waves** |  |  |  |  |
| N1 | 8.2 (0.43 SW/min) | ± 6.2 | 7.0 (0.31 SW/min) | ± 6.1 |
| N2 | 239.2 (1.54 SW/min) | ± 92.6 | 206.1 (1.29 SW/min) | ± 85.1 |
| N3 | 593.9 (4.57 SW/min) | ± 133.8 | 642.5 (4.47 SW/min) | ± 101.9 |
| NREM | 841.4 (2.73 SW/min) | ± 87.5 | 855.6 (2.65 SW/min) | ± 95.5 |
| **Type 2 slow waves** |  |  |  |  |
| N1 | 28.5 (1.35 SW/min) | ± 17.6 | 36.0 (1.63 SW/min) | ± 21.3 |
| N2 | 328.8 (2.09 SW/min) | ± 80.3 | 324.1 (2.06 SW/min) | ± 74.9 |
| N3 | 483.9 (3.75 SW/min) | ± 63.1 | 494.8 (3.44 SW/min) | ± 84.6 |
| NREM | 841.2 (2.73 SW/min) | ± 87.4 | 854.9 (2.65 SW/min) | ± 95.2 |

**Supplementary Table S1 | Raw event counts and densities of sleep spindles and slow waves by sleep stage**

Total number of detected events (N) and corresponding density (events/min, in parentheses) for spindles (all, slow <13 Hz, fast ≥13 Hz), detected on the Fcz derivation, and for slow waves (all, Type 1, Type 2), detected on the signal envelope, separately for N1, N2, N3, and whole NREM sleep, in DOA patients and controls. Values are expressed as mean ± standard deviation across the group.

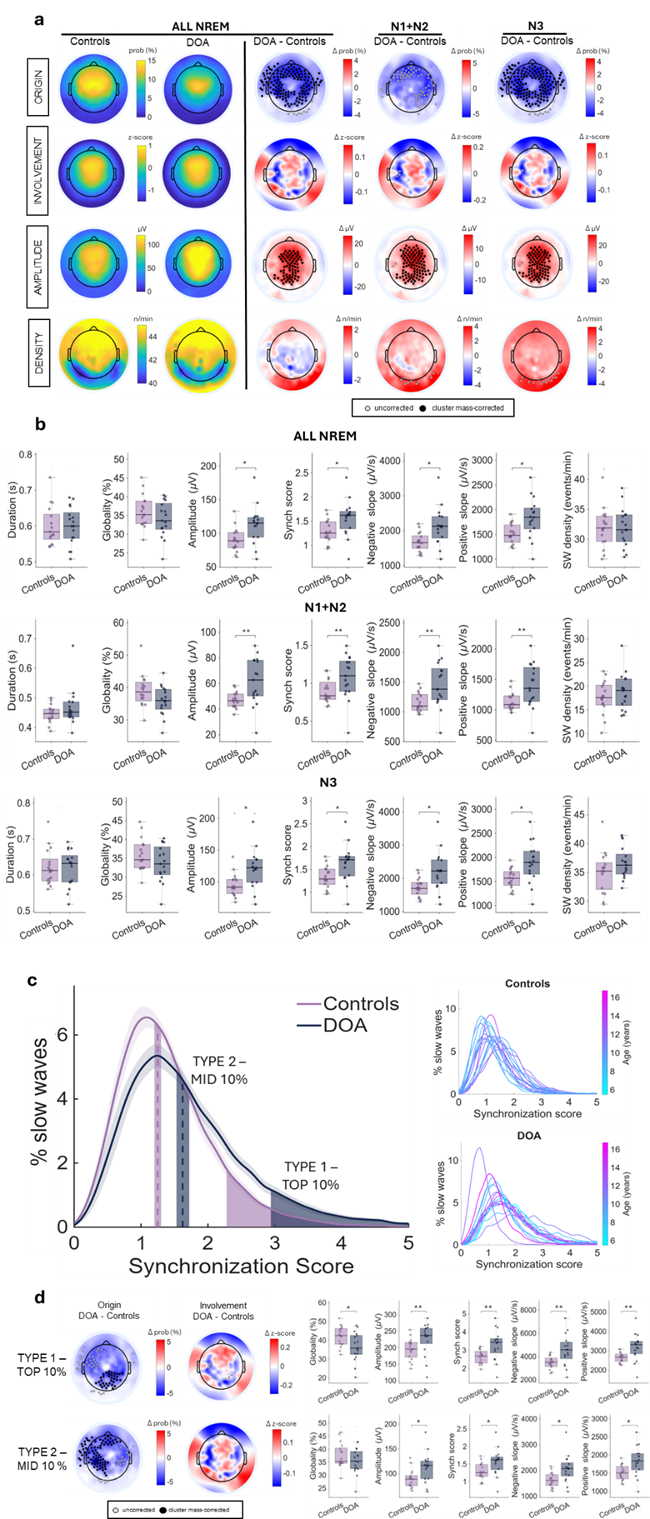

**Supplementary Figure S1 | DOA vs Controls slow-wave characteristics and subtypes during the first hour of NREM sleep**

Replication of the analyses shown in Figure 3 and Figure 4, restricted to the first hour of NREM sleep, when parasomnia episodes preferentially occur and homeostatic sleep pressure is highest. (a) Topographic maps of origin probability, involvement, amplitude and density for controls and DOA (left) and their between-group contrast (DOA - Controls), shown separately for all NREM sleep, N1+N2 and N3. (b) Corresponding boxplots of duration, globality, amplitude, synchronization score, negative and positive slope, and slow-wave density, by sleep stage. (c) Distribution of slow-wave synchronization scores in the two groups (left), with the percentile-based thresholds defining Type 1 (>90th percentile) and Type 2 (45th–55th percentile) waves, and individual age-coded distributions (right). (d) Between-group differences (DOA - Controls) in origin, involvement, globality, amplitude, synchronization score and slopes, separately for Type 1 (top) and Type 2 (bottom) waves. Black dots indicate electrodes within significant clusters (cluster mass-corrected p < 0.05); grey dots show uncorrected channels (p < 0.05). Results in this restricted time window are largely consistent with those obtained across all NREM sleep, with group differences additionally emerging for envelope-identified slow-wave properties in N3 sleep and for Type 2 waves. *q < 0.05; **q < 0.01; ***q < 0.001.
